# Patient Observational Pain and Activity Survey (POPAS) Study using Sequential Electrical Stimulator with Intersperse® Technology in Chronic Axial Spine and Peripheral Joint Pain

**DOI:** 10.1101/2025.06.06.25329103

**Authors:** David Majors, Edward J. Chang, Gilbert Hernandez, Matthew Ibrahim, Sujin Lee

## Abstract

**Objective:** Chronic spine and joint pain are disabling conditions that are prevalent within the general population, but non-invasive and non-pharmacologic treatment options are limited. Sequential Electrical Stimulation (SES) with intersperse therapy is a novel non-invasive, non-pharmacologic technology utilizing a combination of high-frequency interferential therapy for pain relief and neuromuscular electrical stimulation (NMES) for muscle rehabilitation. This study aims to determine the real-world effectiveness of the SES with intersperse in the general population suffering from chronic spine and joint pain.

**Design:** We conducted a cohort study on general population participants with patient observational pain and activity surveys completed upon use of the SES with intersperse device (RS-4i^®^ Plus^TM^) at 1-month, 3-month, and 6-month intervals.

**Results:** The study results showed that among the 61 participants across all treatment groups, the device produced significantly reduced NRS pain scores and improved functional and activity outcomes. The results were consistent in short-term intervals and improved with long-term intervals.

**Conclusion:** The novel SES with intersperse device can significantly reduce pain and improve the daily function of patients in the general population over a long treatment interval. This provides an effective non-invasive and non-pharmacologic treatment for chronic spine and joint pain.

## Introduction

Spine and joint pain are highly prevalent, disabling conditions that have societal and economic ramifications that extend beyond the individual (1). Therefore, it is important to identify comprehensive, effective treatment modalities that can help cut costs to the healthcare system while also improving the quality of life of these individuals (2,3). These conditions are often approached first with conservative, non-invasive methods such as medications, physical therapy, supportive orthotics, and electrical muscle stimulation (4-6).

Classically, muscle stimulation has been seen as a form of neuromuscular electrical stimulation (NMES) which reduces disuse atrophy, reduces muscle spasms and increases circulation to assist in removing metabolites (7). Another type of electric stimulation called high frequency (5000 Hz) interferential therapy decreases the simulation of cutaneous sensory nerves close to the electrodes while penetrating and increasing the effect on deeper tissue (8). Several theories have been proposed to explain the analgesic effects of this therapy including the gate control theory of pain, activation of descending pain suppression pathways, and physiological pain signal blockade (9). The gate control theory of pain involves activating large-diameter A-beta fibers that impede the transmission of pain signals to the brain on small-diameter axons. The activation of descending pain suppression pathways involves the release of endogenous opioids from the rostral ventral medulla and periaqueductal grey matter which help control the nociceptive messages from primary afferent nerves. Lastly, the physiologic pain signal blockade also known as Wedensky inhibition states that the higher frequency prevents C and A-delta fibers from conducting nociceptive impulses (10).

Several studies have demonstrated that when interferential stimulation is used before NMES in a pattern termed sequential electrical stimulation (SES), it provides effective pain control prior to using muscle stimulation and helps to achieve a stronger, more functional contraction without discomfort (11-12). Therefore, development of a novel device that combines both waveforms and provides SES was developed. While the role and effectiveness of SES in treating various chronic pain conditions is still being defined, the goal of our study was to investigate the real-word effectiveness of the SES device in treating musculoskeletal axial and joint pain in the general population as well as its secondary effects on functional outcomes.

## Materials and Methods

### Ethical Statement

The study was approved by the University of California at Irvine Institutional Review Board (UCI IRB) and every participant signed an informed consent prior to participation in the research study. Data obtained during the study is held in the University of California at Irvine RedCap database. This clinical trial was registered with clinicaltrials.gov (Registry # NCT05478265)

### Treatment

The device (RS-4i^®^ Plus^TM^, RS Medical, Vancouver, Washington, USA) is a non-invasive SES system that delivers two waveforms. The first waveform is a pre-modulated, high frequency (5,000 Hz) interferential sine wave across four channels. The second waveform is a low frequency (71 Hz), biphasic, asymmetrical waveform intended for NMES by producing maximal contraction in all muscle types. The device alternates between the two waveforms (sequential electrical stimulation) in a single, uninterrupted treatment called Intersperse™.

### Respondents

The principal investigator and the University of California at Irvine Center for Clinical Research (UCI CCR) partnered with University of California at Irvine (UCI) Marketing to help recruit respondents and potential participants for the study. Direct Communication via university-wide emails, patient clinic encounters and pamphlets distributed in the community were focused areas to recruit respondents. Patient respondents directly communicated with the UCI CCR and principal investigator to gain information about the study. All interested respondents then made an in-person appointment for initial screening with the principal investigator. Any patients that enrolled in the study were prescribed a device by the principal investigator and initial training on use of the device was done by UCI CCR representative at the time of enrollment in the study with receipt of the device through overnight delivery to the participant’s home. The study participants were instructed to use the device up to two sessions per day. Each session consisted of device use at a standard preset program in order to ensure consistency of device program application for each participant. Each session lasted 25 minutes. Each participant was encouraged to bring the device to each follow assessment interval in case device use clarification was needed. The patient respondents were followed throughout the entirety of the six-month long study for their response in use of the device.

### Study Design

All respondents scheduled an initial meeting with the principal investigator to gain information about the study, review the IRB approved protocol, determine eligibility for the study based on criteria, and review the informed consent prior to formally enrolling the in the study. For all participants enrolled, they received tutorial, by a UCI CCR representative, regarding proper pad placement and device use instructions. The inclusion criteria were the participants ages between 18-89, willingness to comply with study protocol, able to sign informed consent form, and have a chronic, persistent musculoskeletal spine pain and/or knee/shoulder joint pain on a numeric rating scale equal or greater than 5 for longer than 12 weeks. There were multiple exclusion criteria including pregnancy, any contraindication to device use, active skin infection at site of use, allergy to electrode adhesive, pacemaker/ICD, abnormal skin sensation, serious psychological disorder, end-stage cardiac/pulmonary/arterial/renal/liver disease, recent vertebral fracture, degenerative/traumatic neuromuscular disease, non-English speaker, uncontrolled diabetes (HbA1c>8.5%), or active cancer. The device was programmed to only have one functioning program to ensure consistency of use during study. Participants were allowed to continue all treatments during study as part of their standard of care for the chronic pain condition. All participants were placed into three groups based on site of chronic pain, such as axial spine, peripheral joint or combination of spine with a joint disease. The groups were further divided into age categories which were ages 65+ and younger than 65. Participants were informed of their rights and were encouraged to report any adverse events to the principal investigator or UCI CCR.

Participant enrollment was completed between January 2023 through December 2023, and all data collection was completed between January 2023 through June 2024. Upon initial prescription of the device, participants completed a baseline assessment after signing the informed consent form. Three follow up assessments were performed at 1 month (30 days +/- 10 days), 3 months (90 days +/- 30 days), and 6 months (180 days +/- 30 days). All baseline and follow up assessments were done with the principal investigator in person or virtual visits based on participant availability. Participants have monthly telephone calls done by UCI CCR representative to help facilitate program adherence with a total of 9 contacts during the six-month study. All participants were given a study ID number and no personal information was collected into UCI RedCap database. All assessments asked for a Numeric Rating Scale (NRS) regarding pain and how pain affects various secondary outcomes. Specifically, the secondary outcomes were activities of daily living, sleeping, lifting, sitting, standing, walking, travelling, muscle spasm, social life, stress, and mood. Upon completion of the six-month study, participants returned the research device and were given a stipend. Participants were also offered a prescription for a replacement device upon return of the research device. Baseline and follow up assessment forms are available as an appendix.

### Data Analysis

We used descriptive statistics to summarize participant’s characteristics, including age group (younger than 65 years old versus 65 or older), previous exposure to TENS therapy, and baseline NRS scores. Baseline scores ranged from 0 – 10, with 0 representing no pain, discomfort, or interference, and 10 representing worst pain or maximum interference. NRS scores were collected for pain before treatment with RS-4i Plus, as well as the degree to which pain interfered with sleep, lifting, sitting, standing, walking and travelling. Additionally, NRS ratings captured pain-related impacts on social life, mood, and stress; and the amount that muscle spasms interfered with daily chores and activities.

For each outcome, we calculated the percent change from baseline to follow-up visits after 1, 3, and 6 months. Given our expectation that treatment would not worsen pain or functional scores, we performed one-sided t-tests to determine whether the reduction in NRS scores at each follow-up visit was significantly greater than zero. We visualized longitudinal trends in NRS outcomes over baseline and 3 follow-up visits using time-series plots. We examined the relationship between the frequency of use of RS-4i Plus and the duration of pain relief after treatments using a heatmap of participant-reported frequency of use by duration of pain relief. Finally, to determine whether any measured patient characteristics were associated with reduced pain scores, we fit a linear fixed-effects model with terms for treatment area (spine, joint, or both), age group (younger than 65 versus 65 or older), previous exposure to TENS therapy, and baseline pain score, and fixed effects for visit number (month 1, 3, or 6) and participant identifier, to account for repeated measures within individuals. We additionally stratified results for changes in NRS pain and function scores by treatment area (spine, joint, or both). We considered hypothesis tests significant at p < 0.05. All analysis was conducted using R version 4.2.1.

## Results

A total of 69 individuals volunteered to undergo the screening process through an initial visit with the principal investigator. During the screening phase, one participant failed to meet eligibility criteria and was excluded, resulting in 68 participants who met all inclusion and exclusion criteria and formally enrolled in the study. Of those enrolled, 7 participants did not complete the study due to withdrawal or lost to follow up. This yielded a final sample of 61 participants who completed the full six-month study period (Figure 1). Participants were categorized into three groups based on the anatomical site of treatment with device. There were 29 participants (47%) in the axial spine only group. There were 11 participants (18%) in the peripheral joint only group. There were 21 participants (35%) in the combination group treating peripheral joint and spine. The majority of the participants (86.9%) were younger than 65 years of age. Due to the small sample size in the older age group, analyses were not stratified by age but rather by treatment location. The prior use of a TENS unit was documented by 49.2% of participants across all groups.

**Figure 1.**
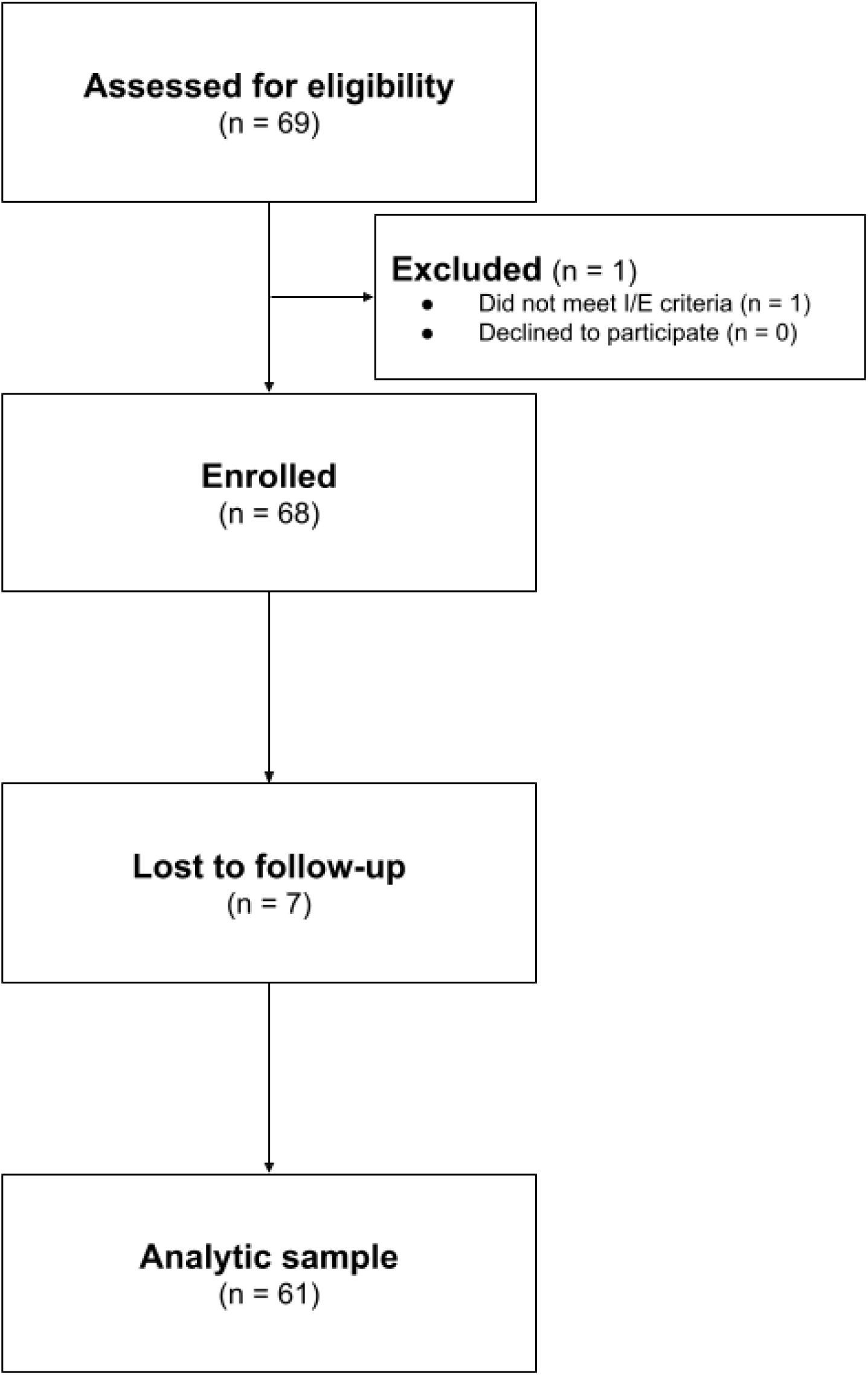
CONSORT diagram displaying sample selection.

Baseline NRS pain scores were higher than 5, as part of the inclusion criteria, and the mean baseline NRS pain scores ranged from 6.7 to 7.2. Baseline NRS scores for functional outcomes in axial group ranged from 4.55 to 7.24. Baseline NRS scores for functional outcomes in the peripheral joint group ranged from 3.36 to 7.27. Baseline NRS scores for functional outcomes in the combination group ranged from 3.76 to 6.76. Overall, the baseline NRS score for functional outcomes across all groups ranged from 4.39 to 6.82 and were generally similar across all groups (Table 1).

**Table 1.** Descriptive summary of the analytic sample and baseline values for all outcomes.

|  | Axial spine | Peripheral Joint | Axial Spine +<br>Peripheral Joint | Overall |
| --- | --- | --- | --- | --- |
| **N (%)** | 29 (47%) | 11 (18%) | 21 (35%) | 61 |
| **Age group (N (%))** |  |  |  |  |
| ≥ 65 | 3 (10.3%) | 0 (0.0%) | 5 (23.8%) | 8 (13.1%) |
| < 65 | 26 (89.7%) | 11 (100.0%) | 16 (76.2%) | 53 (86.9%) |
| **Previous exposure to TENS (N (%))** |  |  |  |  |
| No | 14 (48.3) | 6 (54.5) | 10 (47.6) | 30 (49.2) |
| Yes | 15 (51.7) | 5 (45.5) | 11 (52.4) | 31 (50.8) |
| **Baseline NRS scores (mean (SD))** |  |  |  |  |
| Pain | 6.72 (1.36) | 7.18 (1.60) | 6.76 (1.45) | 6.82 (1.42) |
| Personal care | 5.41 (2.88) | 6.64 (2.11) | 5.67 (2.73) | 5.72 (2.70) |
| Sleep | 6.21 (3.40) | 5.73 (2.80) | 5.19 (2.80) | 5.77 (3.08) |
| Lifting | 7.24 (2.80) | 7.27 (2.80) | 5.95 (3.17) | 6.80 (2.95) |
| Sitting | 5.86 (2.46) | 3.36 (3.50) | 4.48 (2.96) | 4.93 (2.95) |
| Standing | 4.90 (3.48) | 3.82 (3.82) | 4.00 (2.93) | 4.39 (3.34) |
| Travel | 5.17 (3.32) | 4.09 (2.98) | 4.81 (3.30) | 4.85 (3.22) |
| Walking | 5.07 (3.36) | 4.18 (3.82) | 3.76 (3.05) | 4.46 (3.34) |
| Social life | 4.55 (3.48) | 4.09 (2.77) | 5.14 (3.57) | 4.67 (3.37) |
| Mood | 5.76 (2.81) | 4.73 (2.76) | 6.33 (2.74) | 5.77 (2.79) |
| Stress | 5.34 (3.24) | 5.00 (2.83) | 6.14 (2.39) | 5.56 (2.89) |
| Muscle spasms | 5.45 (3.76) | 3.45 (3.67) | 5.52 (3.43) | 5.11 (3.66) |

All NRS pain score reductions were statistically significant (p<0.001) across all treatment groups at all three follow up intervals (Table 2 and Figure 2). The mean NRS pain score reduction show a larger reduction with longer use of the device: 43.4% at 1-month, 61.8% at 3-months, 76.5% at 6-months. Similarly, the NRS scores for functional outcomes showed significant reduction across all strata, again showing improvement in functional outcomes with longer use of the device in the study participants (Table 2 and Figure 3). In heatmaps showing the association between frequency of use of the RS-4i Plus device and duration of pain relief, there was a general trend toward longer-duration pain relief among those using the device more often; the majority receiving more than eight hours of pain relief per treatment (Figure 4). In a linear model, there were no significant associations between the treatment area, age group, or prior TENS exposure and the percentage reduction in pain scores or functional outcome scores from baseline to 1-month, 3-month, and 6-month follow up visits.

**Table 1.** Summary of percent change from baseline to months 1, 3, and 6, stratified by treatment area.

| Follow-up month | Axial Spine |  |  | Peripheral Joint |  |  | Axial Spine + Peripheral Joint |  |  |
| --- | --- | --- | --- | --- | --- | --- | --- | --- | --- |
|  | 1 | 3 | 6 | 1 | 3 | 6 | 1 | 3 | 6 |
| Pain | -43.2%*** | -62.8%*** | -73.0%*** | -49.2%*** | -67.7%*** | -83.2%*** | -37.8%*** | -54.9%*** | -73.3%*** |
| Personal care | -47.1%*** | -65.1%*** | -76.7%*** | -51.1%*** | -71.6%*** | -81.9%*** | -40.6%*** | -60.9%*** | -74.4%*** |
| Sleep | -62.9%*** | -81.6%*** | -89.6%*** | -52.1%*** | -82.9%*** | -89.6%*** | -35.3%*** | -57.4%*** | -80.3%*** |
| Lifting | -44.2%*** | -63.5%*** | -71.1%*** | -45.0%** | -56.7%*** | -78.0%*** | -37.3%*** | -57.8%*** | -79.5%*** |
| Sitting | -50.8%*** | -73.0%*** | -82.3%*** | -52.0%** | -67.9%*** | -91.7%*** | -36.5%*** | -56.0%*** | -70.0%*** |
| Standing | -48.9%*** | -65.0%*** | -74.1%*** | -39.0%* | -59.0%** | -81.2%*** | -49.7%*** | -68.2%*** | -83.9%*** |
| Travel | -49.8%*** | -65.0%*** | -72.7%*** | -58.5%*** | -78.1%*** | -97.8%*** | -42.1%*** | -72.9%*** | -86.6%*** |
| Walking | -56.1%*** | -70.7%*** | -77.8%*** | -70.1%*** | -78.3%*** | -94.6%*** | -53.6%*** | -72.0%*** | -85.3%*** |
| Social life | -53.5%*** | -69.6%*** | -77.8%*** | -49.4%*** | -66.3%*** | -87.3%*** | -37.5%*** | -66.8%*** | -82.9%*** |
| Mood | -55.9%*** | -71.0%*** | -79.4%*** | -46.4%** | -60.2%*** | -82.9%*** | -35.1%*** | -57.2%*** | -77.3%*** |
| Stress | -46.7%*** | -65.6%*** | -80.6%*** | -54.7%*** | -65.9%*** | -85.0%*** | -27.2%*** | -51.4%*** | -72.3%*** |
| Muscle spasms | -51.1%*** | -78.6%*** | -87.1%*** | -70.1%** | -75.7%*** | -83.3%*** | -52.0%*** | -66.3%*** | -84.2%*** |

Percent change from baseline calculated using all observations with non-zero baseline values.
\* $p < 0.05$ ; \*\* $p < 0.01$ ; \*\*\* $p < 0.001$ .

**Figure 2.**
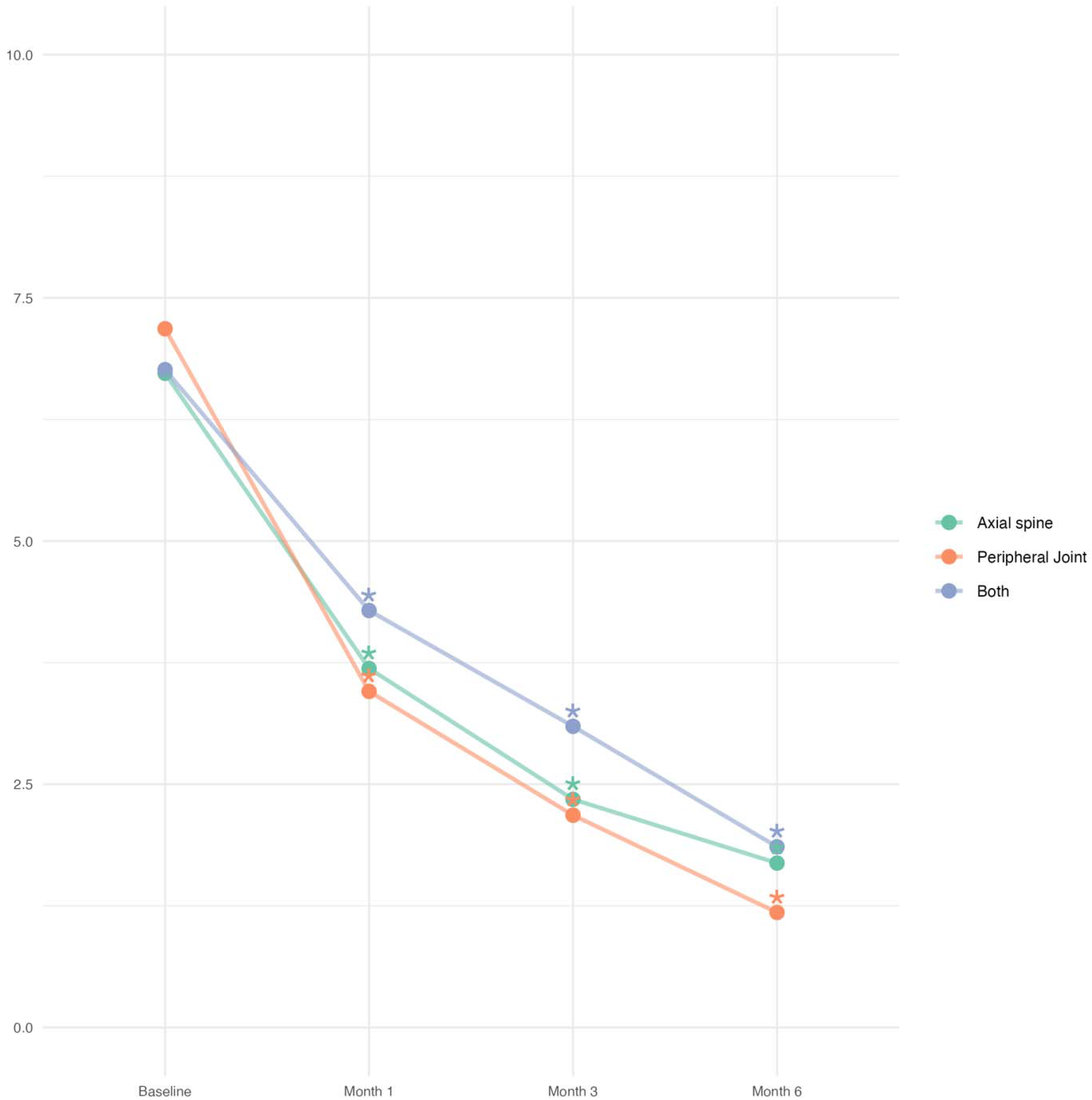
Time-series plot showing change in NRS pain scores from baseline visit and month 1, month 3, and month 6 follow-up visits. * indicates significant difference from baseline value at p < 0.05.

**Figure 3.**
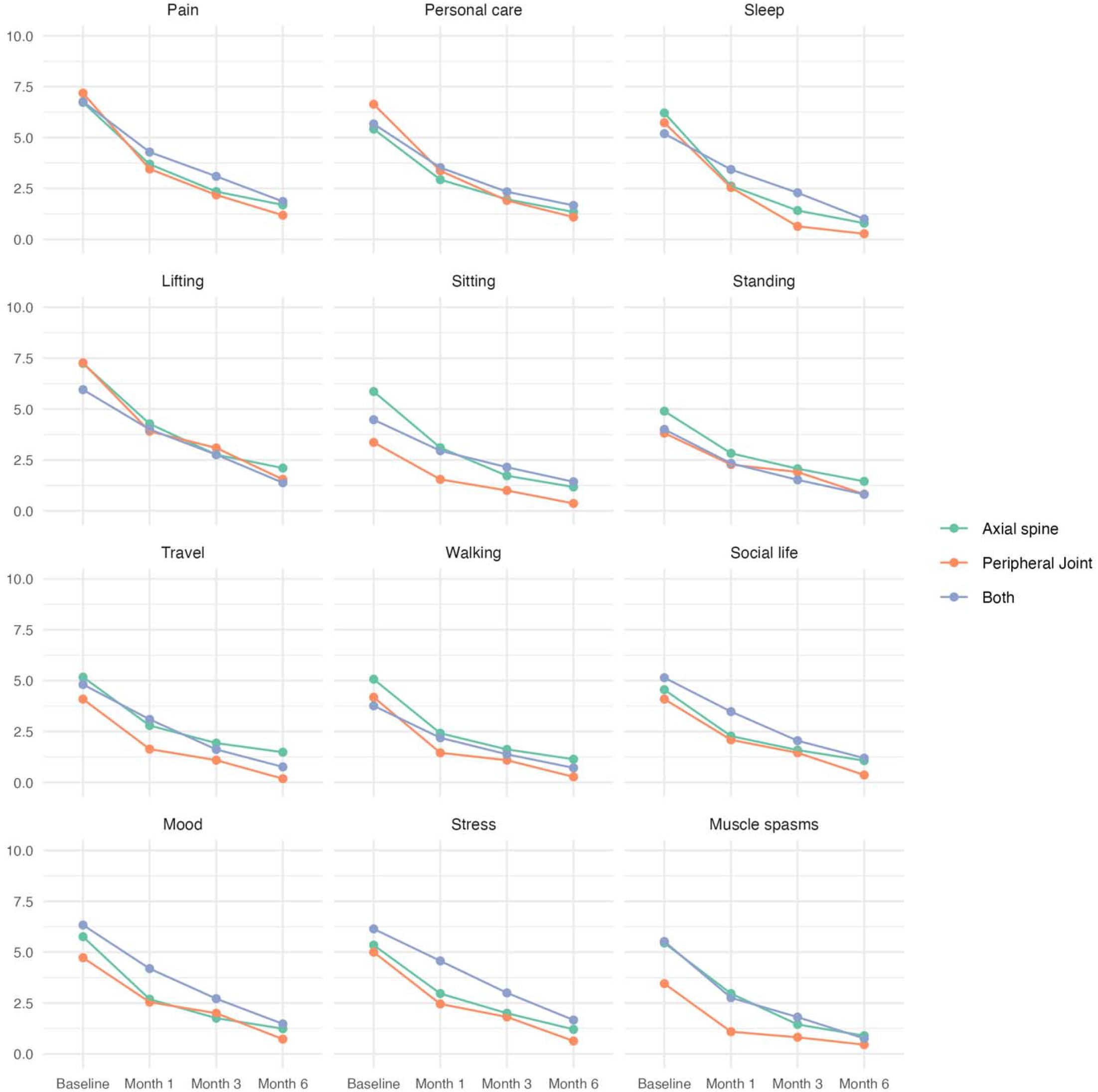
Time-series plots showing NRS scores for all outcomes at baseline and at month 1, month 3, and month 6 follow-up visits.

**Figure 4.**
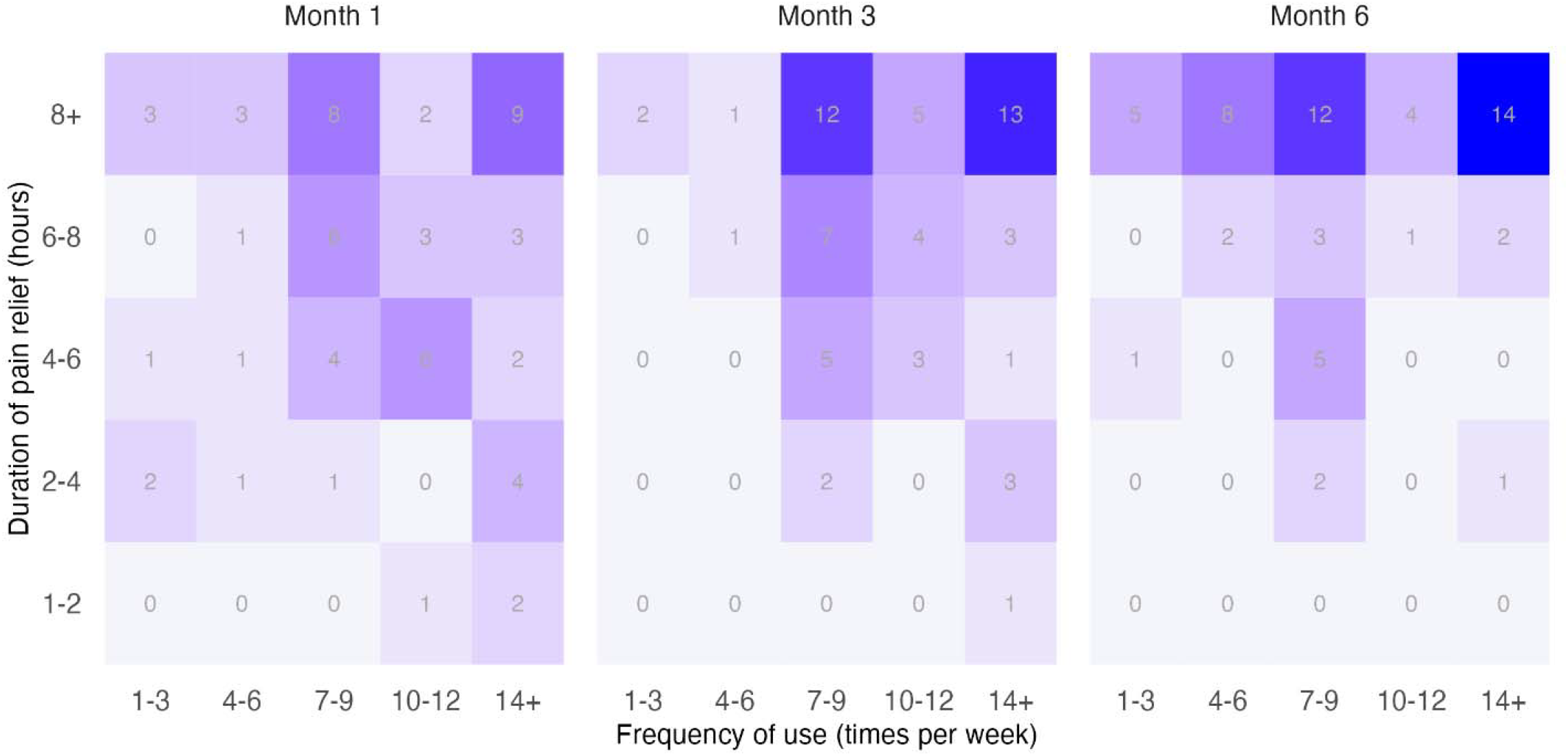
Heatmap showing the association between frequency of use of the RS-4i Plus device and duration of pain relief.

At the final follow up visit, 56 of the 61 participants (91.8%) elected to continue using the RS-4i Plus neuromodulation device and accepted a prescription to support continuity of care beyond the study period. Additionally, the majority of participants (73.8%) reported continued daily use of the device at the time of the final visit.

## Discussion

In this prospective study, we evaluated the effectiveness of a novel sequential electrical stimulation (SES) device with Intersperse^®^ technology in managing musculoskeletal pain of the spine and joints. Our findings demonstrate consistent and statistically significant reduction in patient-reported pain and functional NRS scores over a six-month period. Improvement in function were observed across multiple quality of life domains, including sleep, mobility, social participation, and mood.

Previous studies have demonstrated the efficacy of individual conservative neuromuscular stimulation modalities in alleviating lumbosacral degenerative pain and osteoarthritic knee pain (13-17). However, to our knowledge, no prior research has examined a combined modality that integrates both interferential therapy and neuromuscular electrical stimulation (NMES) through Intersperse^®^ signal delivery. This technology takes advantage of the quiescent phases of NMES cycle by interjecting interferential stimulation that targets larger diameter A-beta fibers via the gate control theory of pain. This mechanism is designed to facilitate effective analgesia and improve tolerance during muscle stimulation (18). This synergistic use of combined waveforms optimizes therapeutic outcomes, such as reduction of treatment duration while minimizing patient discomfort. Our study aimed to evaluate the effects of this novel stimulation technique on pain, incorporating longitudinal outcome across three follow-up intervals; 1-month, 3-months, and 6-months. This design allowed for a more comprehensive assessment of the therapy’s clinical utility over time in a general outpatient population.

Our findings demonstrated a statistically significant reduction in both pain and functional NRS scores from baseline at all follow-up intervals (p<0.001), without significant differences in outcomes based on treatment region or age group (Table 2). These results support broad applicability of the SES with intersperse device across a general outpatient population. Each follow up assessment displayed a reduction in pain by 43.4% at the 1-month interval and 76.5% at the conclusion of the study. This reduction of NRS pain scores correlated to improvements in functional secondary outcomes, including activities of daily living (ADLs), sleep, lifting, sitting, standing, walking, travel capability, muscle spasms, mood, stress, and social engagement. No adverse events were attributed to use of the SES with intersperse device, suggesting it is both safe to use and well-tolerated for daily use in the general population.

While exploratory, heatmap analysis showed a trend toward longer duration of pain relief among participants who used the device more frequently. At the final follow-up visit, 91.8% of participants opted to continue using the device and accepted a prescription, with the majority still using the device on a daily basis. These findings suggest that the RS-4i Plus SES with Intersperse^®^ device may serve as a meaningful conservative option for patients seeking relief from chronic axial or joint pain, particularly those for whom pharmacologic therapies are limited by side effects or invasive treatments are contraindicated. This device offers a potentially durable, low risk alternative for managing chronic musculoskeletal pain.

Despite these promising results, this study has several limitations. The primary limitation is the lack of a control group. Given the nature of this prospective study and the use of a prescribed neuromodulation device, we anticipated that participant adherence would be challenging if a placebo device were used, particularly in individuals experiencing chronic pain. As a result, a control group was not included.

Secondarily, the outcomes relied on subjective self-reported NRS scores, which may be influenced by individual variability in pain perception, recall bias, and psychological factors. In addition, the participants continued to receive the standard of care treatment for their chronic pain from their respective medical providers during the study period. This introduces variability in pain management approaches that may have impacted individual outcomes.

Future investigations should prioritize patients with uncontrolled diabetes typically defined as a hemoglobin a1c > 7% as these individuals are often excluded from receiving corticosteroid injections due to potential complications from glycemic variability (19). Also, randomized controlled trials comparing SES therapy to a control or placebo group, where feasible, would provide more robust evidence for its efficacy. Finally, incorporation of objective functional measures and pain-related biomarkers could further strengthen the findings.

## Conclusion

In this prospective observational study, the use of the RS-4i^®^ Plus Sequential Stimulator with Intersperse^®^ technology was associated with significant and sustained improvements in both pain and functional outcomes among individuals with axial spine and peripheral joint pain. Participants reported meaningful reductions in pain interference across multiple domains of daily life, with benefits observed regardless of age or treatment area.

These findings support the utility of the sequential electrical stimulation with intersperse device as a conservative, non-pharmacologic option for managing chronic musculoskeletal pain. Although limitations such as lack of a control group and reliance on self-reported outcomes must be acknowledged, the results are promising and suggest the SES with intersperse device use may be as a safe and effective non-interventional treatment option for individuals with challenging chronic pain. Future research should explore the role of SES in more diverse populations, including those with limited access to pharmacologic or interventional treatment options, such as individuals with poorly controlled diabetes, to expand understanding of the device’s broader applicability.

## Acknowledgments

The authors thank the support of University of California at Irvine Center for Clinical Research and Kyle Hart for data processing.

## Data Availability Statement

The datasets generated during and/or analyzed during the current study are available from the corresponding author on reasonable request.

